# CUSUM Analysis of the Initial Learning Curve for Robot-Assisted Gynecologic Surgery at a German Tertiary Center

**DOI:** 10.64898/2026.07.13.26357950

**Authors:** Morva Tahmasbi Rad, Dario Colacurci, Elias Bascharyar, Giuseppe Bifulco, Sven Becker, Ina Shehaj

## Abstract

**Objective:** To evaluate the initial learning curve of robot-assisted laparoscopic hysterectomy performed by a single surgeon during the implementation of robotic gynecologic surgery at a German tertiary university center using cumulative sum (CUSUM) analysis.

**Methods:** This retrospective observational study included the first 66 consecutive patients who underwent robot-assisted laparoscopic hysterectomy using the da Vinci X surgical system between January 2022 and September 2023 at the University Hospital Frankfurt. Operative performance was assessed using CUSUM analysis of skin-to-closure time, console time, and docking time. Based on the CUSUM curve for operative time, cases were divided into two phases: phase 1 (cases 1–36) and phase 2 (cases 37–66). Demographic, perioperative, and postoperative outcomes were compared between phases.

**Results:** Sixty-six patients were included. Median age was 48.5 years, and median BMI was 27.45 kg/m². Previous abdominal surgery was present in 80.3% of patients, and 47% had endometriosis. At least 25% of cases had elevated BMI and large uterine volume. CUSUM analysis identified a transition point after 36 cases, indicating completion of the initial learning phase. Median skin-to-closure time significantly decreased from 108 minutes in phase 1 to 85.5 minutes in phase 2 (p=0.029). Console time and docking time showed progressive improvement, although these differences did not reach statistical significance. Perioperative outcomes, complication rates, conversion rates, postoperative pain scores, and hospital stay were comparable between phases.

**Conclusion:** Robot-assisted laparoscopic hysterectomy demonstrates a well-defined learning curve, with procedural stabilization achieved after approximately 36 consecutive cases. The successful and safe implementation of robotic gynecologic surgery, even for complex cases, is feasible during the initial adoption phase at a tertiary university center when supported by structured training and standardized workflows.

## 1. Introduction

Robotic-assisted surgery (RAS) is considered an increasingly important component of modern gynecologic practice. Initially developed to overcome technical limitations of conventional minimally invasive surgery, robotic technology has progressively expanded across multiple surgical fields and gynecologic indications, including hysterectomy, lymphadenectomy, myomectomy, sacrocolpopexy, and endometriosis surgery [1–3]. The technical advantages of RAS include three-dimensional visualization, tremor filtration, improved dexterity, articulated instruments, and improved surgeon ergonomics. These characteristics may facilitate complex dissection and intracorporeal suturing, particularly in high-complex pelvic procedures where conventional laparoscopy may become challenging [2–4]. RAS procedures have been associated with perioperative benefits when compared with open surgery, such as reduced blood loss, shorter hospital stay, and lower postoperative morbidity [1,2]. In gynecology, robotic surgery has shown capacity in complex surgical settings and high-risk patients; in this context, improved instrument control and higher visualization may facilitate minimally invasive approaches [4,5]. However, RAS superiority over traditional laparoscopy is still debated. Current evidence suggests broadly comparable perioperative outcomes in routine benign and oncologic gynecologic procedures, although robotic surgery is often associated with longer operative times and higher institutional costs [3,6]. Therefore, the clinically relevant question is not whether robotic surgery is universally superior to laparoscopy, but under which technical and organizational conditions it provides meaningful added value. One of the most important factors of successful robotic implementation is the learning curve. As with any new surgical technology, the adoption phase needs progressive gain of technical competence, workflow optimization, and effective coordination of the operating room team. Ensuring safe implementation requires objective evaluation of this process. Indeed, previous studies have suggested that RAS may facilitate acquisition of minimally invasive surgical skills and simplify technical ability when compared with conventional laparoscopy [7–9]. Similar observations have been reported in gynecologic minimally invasive surgery, where operative performance improves significantly with procedural experience [10]. Cumulative sum (CUSUM) analysis is a consistent statistical method for evaluating surgical learning curves; it is based on monitoring performance and identifying transition points between early learning and procedural stabilization [7,8]. This methodology has proven applicable for assessing operative efficiency and technical progression during implementation of new surgical platforms, including robotic gynecologic surgery [11]. The present study analyzes the initial experience of implementing robot-assisted gynecologic surgery using the da Vinci X system at a tertiary university center in Frankfurt, Germany. The aim was to evaluate the learning curve of a single surgeon during the first consecutive robotic-assisted laparoscopic hysterectomies, with particular focus on skin-to-closure time, console time, docking time, perioperative outcomes, and training implications.

## 2. Material and Methods

### 2.1 Study design and ethical approval

This is a retrospective observational study conducted at the Department of Obstetrics and Gynecology, University Hospital Frankfurt, Germany. The study included the first 66 consecutive patients who underwent robot-assisted laparoscopic hysterectomy using the da Vinci X Surgical System (Intuitive Surgical Inc., Sunnyvale, CA, USA) between January 2022 and September 2023. All procedures were performed by the same surgeon (M.T.R.) as the primary console surgeon. The analyzed cohort therefore represents the initial robotic experience of a single operator and the associated operating room team, both without prior independent or dependent robotic surgical experience. No case involved console sharing with another surgeon, and no additional robotic console operator participated during the analyzed procedures. Before implementation of the robotic program, the surgeon and operating room staff completed the standard manufacturer-certified training pathway, including simulation-based instruction, system familiarization, and observational proctoring. Ethical approval was obtained from the institutional Ethics Committee of Goethe University; Frankfurt-Germany (approval no. 2023-1476) in accordance with the ethical standards of the 1964 Declaration of Helsinki and its later amendments. This approval specifically authorized the retrospective identification and pseudonymized analysis of clinical data for patients. Written informed consent was obtained from all patients during follow-up assessment. The clinical management of the entire study population adhered to established national guidelines and institutional protocols.

### 2.2 Surgical procedures and data collection

All procedures were performed using a three-arm da Vinci X robotic platform. Before implementation of the robotic program, the surgeon and surgical staff completed the standard manufacturer-certified training pathway, including simulation-based instruction, system familiarization, and observational proctoring. The main aims during this phase were minimization of conversion and perioperative complications, and optimization of operative workflow. Following completion of the initial learning phase, more complex cases, including advanced endometriosis and gynecologic oncologic procedures, were progressively introduced. For benign indications, total hysterectomy with bilateral salpingectomy or salpingo-oophorectomy was performed. In malignant cases, pelvic sentinel lymph node biopsy was additionally performed when indicated. Clinical and perioperative data were retrospectively extracted from institutional medical records and the Intuitive surgical database. Collected variables included: Age, body mass index (BMI), menopausal status, previous abdominal surgery, previous cesarean section, uterine volume, adhesion score, American Society of Anesthesiologists (ASA) physical status classification, skin-to-closure time, console time, docking time, estimated blood loss, conversion rate, postoperative pain scores, length of hospital stay, and perioperative complications.

### 2.3 Definitions and classifications

Intraoperative adhesions were graded according to the Zühlke classification [12]. Patient preoperative physical status was classified according to the American Society of Anesthesiologists (ASA) Physical Status Classification System [13]. Postoperative pain was assessed using the 11-point Numeric Rating Scale (NRS), ranging from 0 (no pain) to 10 (worst imaginable pain), which is a validated and widely accepted instrument for pain assessment [14].

Docking time was defined as the interval between positioning of the robotic cart at the operating table and complete connection of robotic arms with insertion of all instruments into the operative field [15]. Operative time was defined as the interval from skin incision to final wound closure. Conversion was defined as inability to complete the planned robotic procedure requiring transition to conventional laparoscopy or laparotomy. Postoperative complications were defined as any deviation from the normal postoperative course occurring within 30 days and were classified according to the Clavien–Dindo system [16].

### 2.4 Outcome measures

The primary endpoint was evaluation of the robotic learning curve through operative efficiency assessment. Learning curve analysis focused on: skin-to-closure time, console time, docking time. Secondary endpoints included perioperative safety outcomes, including blood loss, conversion rate, postoperative complications, postoperative pain scores, and length of hospital stay.

### 2.5 Statistical analysis

The learning curve was evaluated using cumulative sum (CUSUM) analysis [17,18]. Cases were arranged chronologically according to operative date. CUSUM values were calculated using the formula:

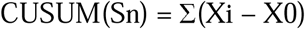

where Xi represents the operative time for each individual case and X0 represents the overall mean operative time of the complete series. CUSUM curves were generated for skin-to-closure time, console time, and docking time to identify inflection points corresponding to transition between learning phases. Based on these inflection points, the cohort was divided into two phases for comparative analysis. Continuous variables are presented as median and interquartile range (IQR); instead, categorical variables are presented as frequencies and percentages. Group comparisons were performed using the Mann–Whitney U test for continuous variables and Pearson’s chi-square test or Fisher’s exact test for categorical variables, as appropriate. A two-sided p-value <0.05 was considered statistically significant. Statistical analyses were performed using SPSS version 26.0 (IBM Corp., Chicago, IL, USA).

## 3. Results

A total of 66 consecutive patients who underwent robot-assisted laparoscopic hysterectomy between January 2022 and September 2023 were included in the analysis. According to CUSUM analysis of skin-to-closure time, the learning process was divided into two distinct phases: Phase 1 (cases 1–36), representing the initial implementation period, and Phase 2 (cases 37–66), reflecting procedural consolidation after attainment of technical proficiency.

### 3.1 Patient characteristics and procedural complexity

Baseline demographic and clinical characteristics are summarized in Table 2. The median patient age was 48.5 years (IQR 43.0–56.3), with no significant difference between the two phases (*p*=0.536). Median BMI was 27.45 kg/m² (IQR 23.9–33.1), with a non-significant trend toward higher BMI in Phase 2 (*p*=0.053); 25% the cases had the BMI over 33 kg/m² (maximum 42.2 kg/m²). Previous abdominal surgery was highly prevalent, reported in 80.3% of patients, while 48.5% were postmenopausal. Endometriosis represented either the primary surgical indication or a concomitant intraoperative diagnosis in 47.0% of cases, highlighting the substantial complexity of the surgical cohort. The median uterine volume was 144.9 mL (IQR 72.8–312.7), without significant intergroup differences (*p*=0.616); 25% of this cohort had the Uterus Volume over 315 ml (to maximum 940ml). Moderate-to-severe adhesions (Zühlke grade >2) were identified in 41.0% of procedures. Most patients (92.4%) were classified as ASA physical status I–II, indicating a generally favorable perioperative risk profile. No statistically significant differences in baseline clinical complexity were observed between the two learning phases, supporting comparability of the cohorts. Table 3 shows the surgical complexity indicators stratified by learning phase.

**Table 1.** Classification of intraoperative adhesions according to the Zühlke adhesion score [12].

| Grade | Observation |
| --- | --- |
| 0 | No adhesions |
| 1 | Filmy adhesions; easily separated by blunt dissection without vascularization |
| 2 | Stronger adhesions; blunt dissection sufficient, with partial sharp dissection possible; beginning vascularization |
| 3 | Strong adhesions requiring sharp dissection; clear vascularization |
| 4 | Very strong adhesions requiring sharp dissection only, with severe organ attachment and increased risk of organ injury |

**Table 2.**
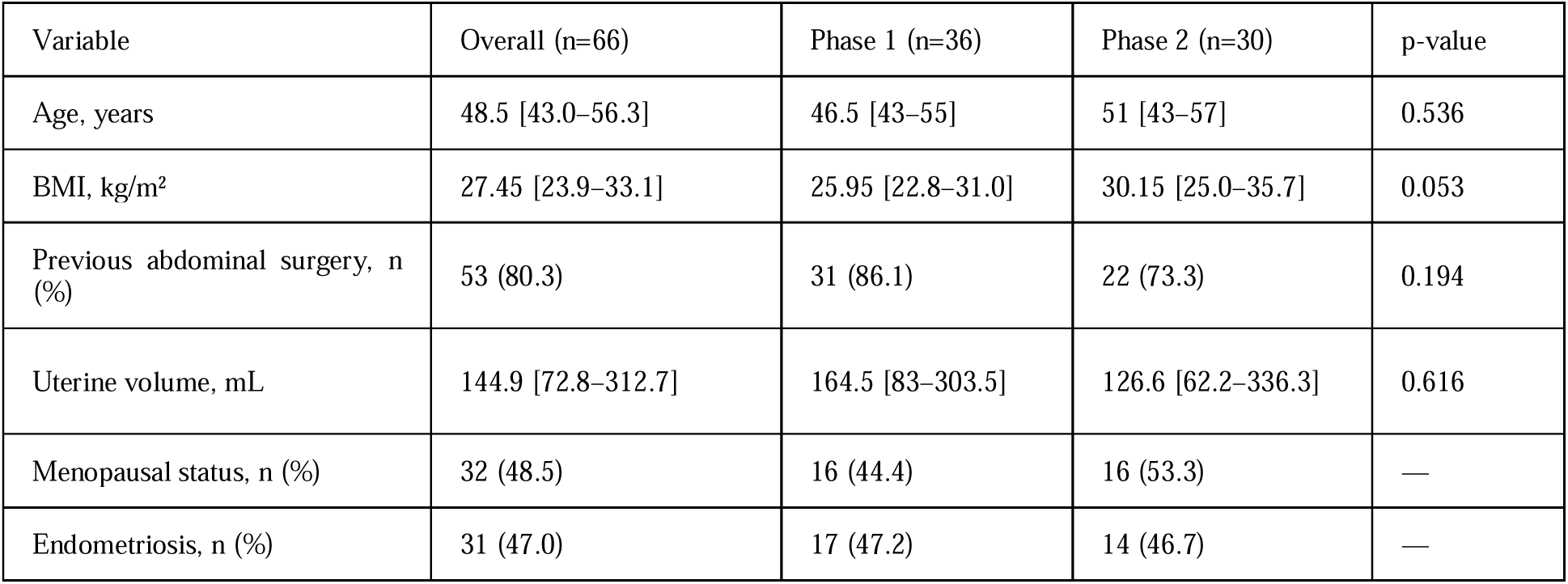
Baseline demographic and clinical characteristics stratified by learning phase.

**Table 3.**
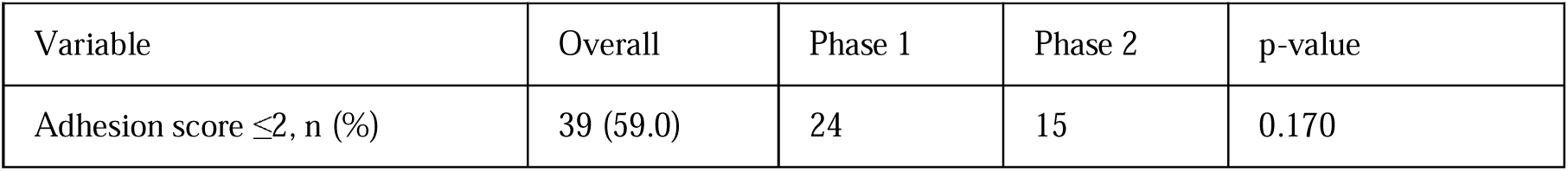
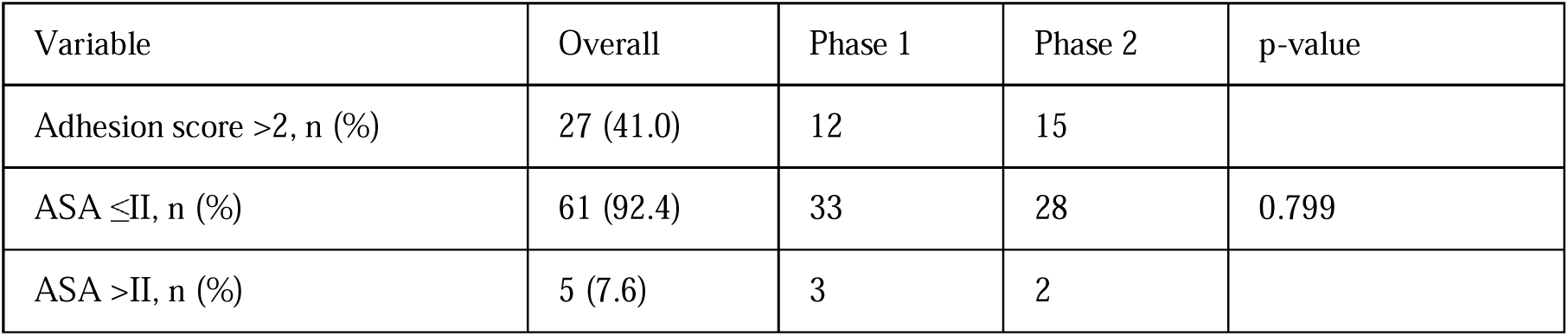
Surgical complexity indicators stratified by learning phase.

| Variable | Overall | Phase 1 | Phase 2 | p-value |
| --- | --- | --- | --- | --- |
| Adhesion score ≤2, n (%) | 39 (59.0) | 24 | 15 | 0.170 |
| Adhesion score >2, n (%) | 27 (41.0) | 12 | 15 |  |
| ASA ≤II, n (%) | 61 (92.4) | 33 | 28 | 0.799 |
| ASA >II, n (%) | 5 (7.6) | 3 | 2 |  |

### 3.2 Operative performance outcomes

Operative performance data are presented in Table 4. A statistically significant reduction in skin-to-closure time was observed between the two phases, decreasing from 108 minutes (IQR 73–128.5) in Phase 1 to 85.5 minutes (IQR 65.8–108) in Phase 2 (*p*=0.029), indicating significant procedural optimization over time. Median console time also decreased from 66.5 to 62 minutes, showing a favorable trend toward improved surgical efficiency, although statistical significance was not reached (*p*=0.074). Similarly, docking time declined from 24 to 18.5 minutes, reflecting progressive optimization of robotic setup and team coordination, although this reduction did not reach statistical significance (*p*=0.168). Notably, active instrument usage time showed borderline statistical improvement (*p*=0.050). Positioning and draping time significantly increased in Phase 2 (p=0.008), a finding plausibly explained by the progressive introduction of more complex cases and the non-significant trend toward higher BMI observed in this phase (Table 2), both of which require more extensive patient positioning and padding. No significant differences were observed in total theater time or anesthesia-related intervals.

**Table 4.**
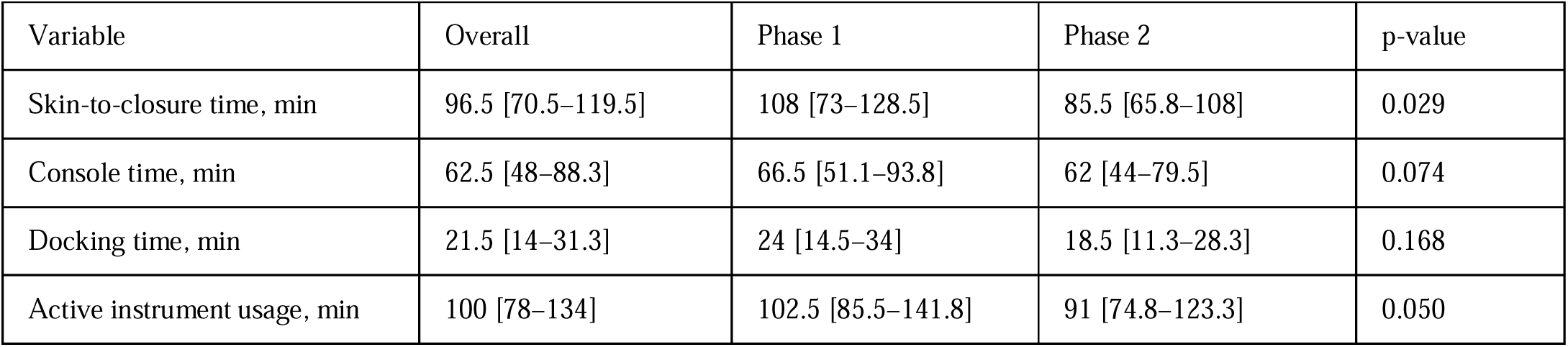
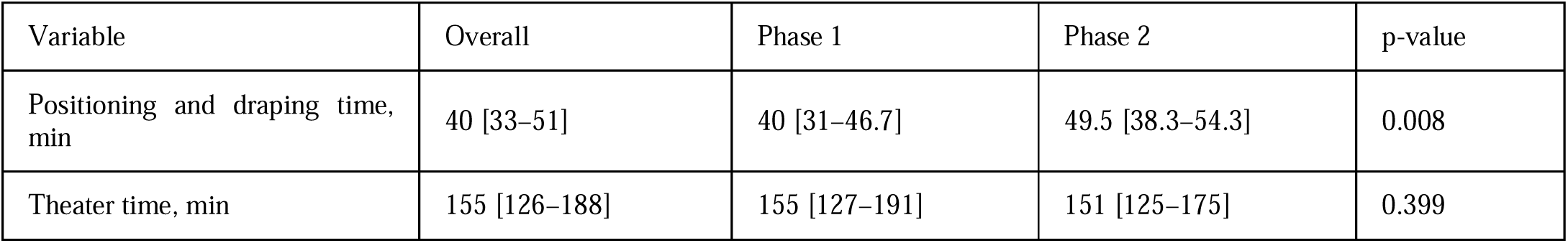
Operative performance metrics stratified by learning phase.

### 3.3 Perioperative safety outcomes

Perioperative safety outcomes are summarized in Table 5. The overall conversion rate was 10.6% (7/66), with no significant difference between learning phases (*p*=0.884). One conversion during Phase 1 was necessitated by severe adhesions with uterine immobility. The remaining conversions in both phases were related to large uterine size associated with intraoperative suspicion of malignancy, prompting avoidance of morcellation. The overall pereioperative complication rate was 7.6%, without significant intergroup differences (*p*=0.497); the complications included 2 cases of wound infection, 2 cases of urinary tract infection and 1 case of pneumothorax during the intubation phase. Median postoperative pain scores remained consistently low both before discharge and after discharge, with no differences between phases. Length of hospitalization and delayed drain removal also remained stable throughout the learning process.

**Table 5.** Perioperative safety outcomes stratified by learning phase.

| Variable | Overall | Phase 1 | Phase 2 | p-value |
| --- | --- | --- | --- | --- |
| Hemoglobin drop, g/dL | -0.8 [-1.48 to -0.3] | -0.8 [-1.6 to -0.5] | -0.6 [-1.3 to -0.25] | 0.286 |
| Pain score before discharge | 3 [2–3] | 3 [2–3] | 3 [2–3] | 0.686 |
| Pain score after discharge | 1 [0–2] | 1 [0.25–2] | 1 [0–1.25] | 0.215 |
| Postoperative complications, n (%) | 5 (7.6) | 2 (5.6) | 3 (10.0) | 0.497 |
| Prolonged hospital stay (>3 days), n (%) | 9 (13.6) | 4 (11.1) | 5 (16.7) | 0.513 |
| Conversion, n (%) | 7 (10.6) | 4 (11.1) | 3 (10.0) | 0.884 |

### 3.4 Learning curve analysis

CUSUM analysis demonstrated a clear biphasic learning pattern across all evaluated operative metrics. The skin-to-closure time CUSUM curve identified a distinct inflection point at case 36, marking the transition from the initial acquisition phase to procedural consolidation (Figure 1). The console time CUSUM curve demonstrated a similar pattern, confirming progressive technical mastery and increasing console efficiency (Figure 2). For docking time, the maximal peak was observed at case 28, followed by a progressive downward trend, reflecting maturation of team coordination and standardization of robotic setup (Figure 3).

**Figure 1.**
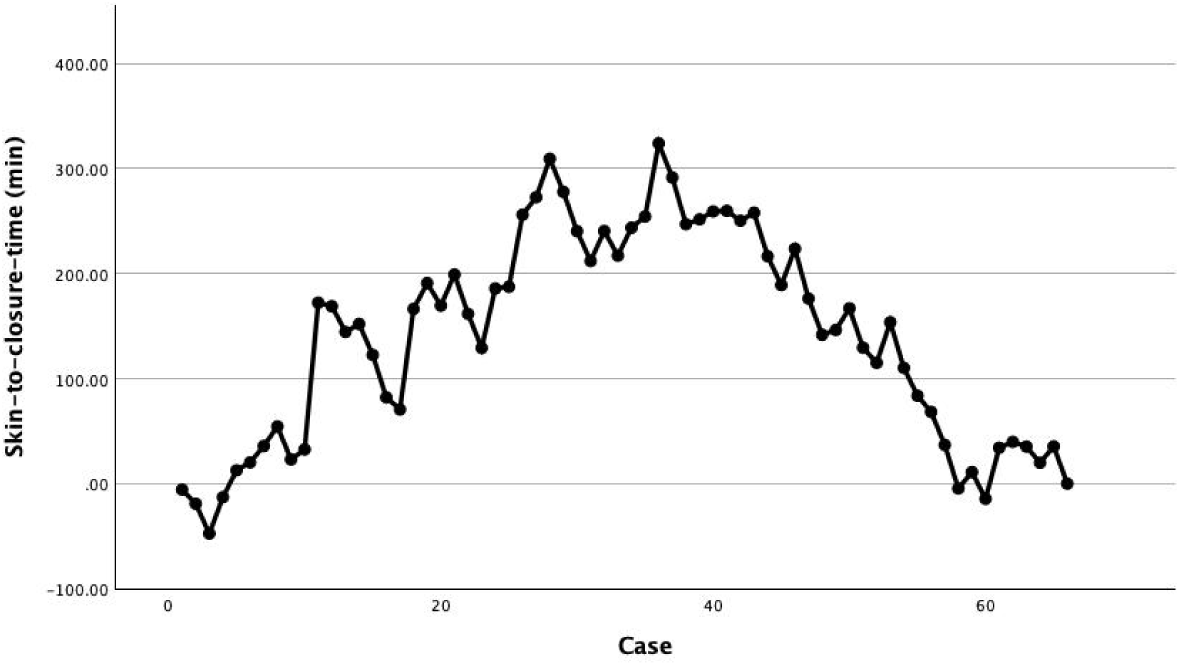
CUSUM curve for skin-to-closure time demonstrating the transition from the initial learning phase to procedural consolidation at case 36.

**Figure 2.**
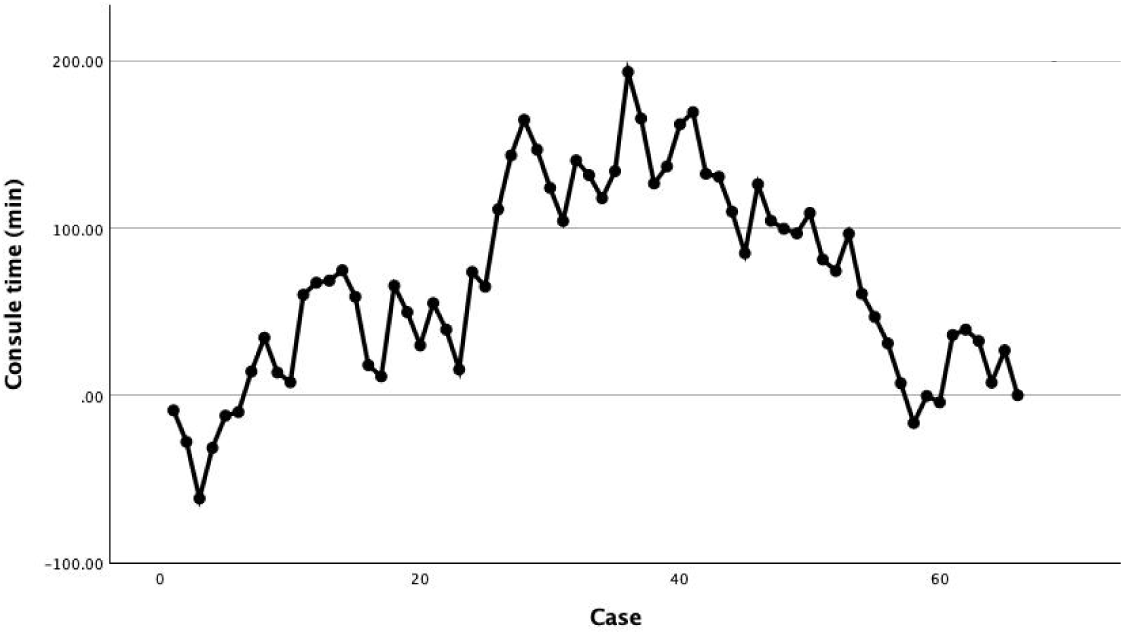
CUSUM curve for console time showing progressive reduction in console-related operative duration.

**Figure 3.**
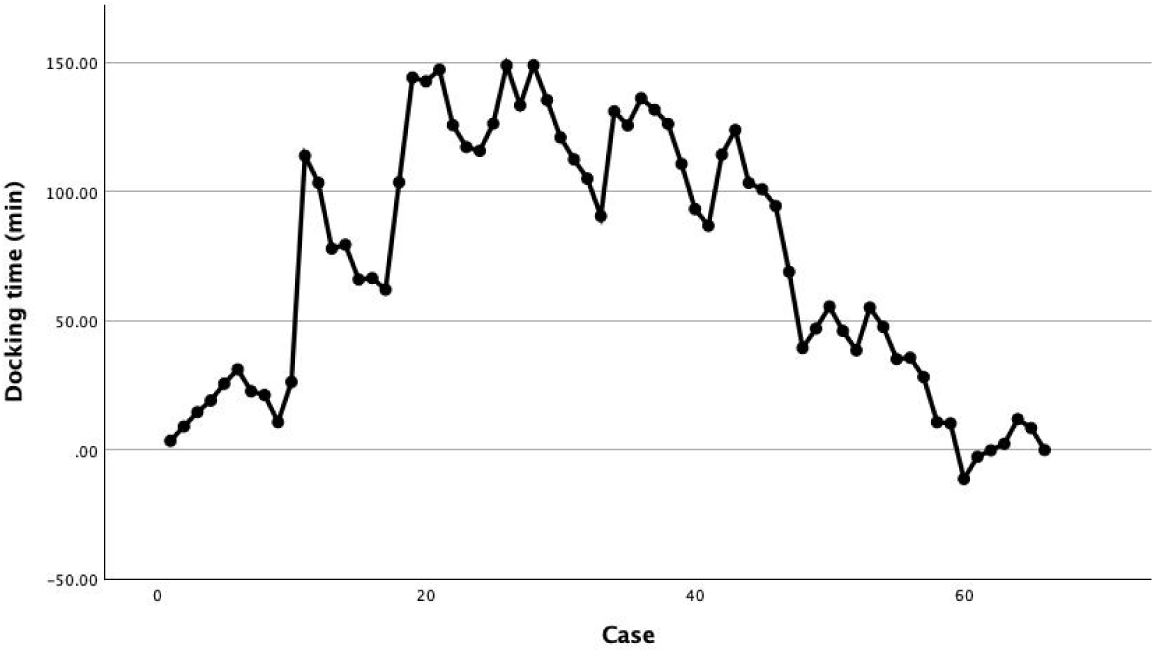
CUSUM curve for docking time, with peak technical adaptation observed at case 28 followed by workflow stabilization.

## 4. Discussion

This study evaluated the initial implementation of robot-assisted laparoscopic hysterectomy in a German tertiary university center. The main finding was the presence of a clear learning curve. CUSUM analysis identified a transition point after 36 cases for skin-to-closure time. After this point, operative efficiency improved significantly, while perioperative safety remained stable. The reduction in skin-to-closure time from 108 minutes in Phase 1 to 85.5 minutes in Phase 2 suggests progressive procedural optimization. Console time and docking time also decreased, although these differences did not reach statistical significance. Indeed, this pattern is clinically relevant: it suggests that the learning process was not limited to the surgeon’s console performance, but also involved the wider robotic workflow, including patient positioning, trocar placement, docking, and team coordination. The observed transition after approximately 36 cases is consistent with previous CUSUM-based analyses of robot-assisted hysterectomy [19–21]. In similar cohorts, procedural proficiency has been reported after approximately 30–35 cases, with reductions in total operative time, console time, docking time, and blood loss after the initial phase. Comparative data between newly introduced robot-assisted hysterectomy and established laparoscopic hysterectomy also suggest that total operative time may become comparable after approximately 30 cases [21], whereas pure operative or console time may improve earlier. These findings support the concept that robotic learning is composed of different components. Surgeon-dependent technical steps may improve relatively early, while complete procedural efficiency requires further experience and workflow standardization. Our cohort included a high proportion of patients with a history of previous abdominal surgery and endometriosis. Furthermore, patients with elevated body mass index (BMI) and large uterine volume were not excluded, and 41% of cases presented with moderate-to-severe intra-abdominal adhesions. Despite this high level of surgical complexity, perioperative outcomes remained favorable, with no significant differences in complication rates, conversion rates, postoperative pain scores, or length of hospital stay across the different phases of the learning curveThis is an important finding, suggesting that the initial adoption of robotic surgery also in complex cases can be safe when case selection is careful and the surgical team follows a structured training pathway [22]. The overall conversion rate was 10.6%. Most conversions were not attributable to technical inability to complete the robotic dissection, but to intraoperative suspicion of malignancy in the presence of an enlarged uterus. In these cases, the decision was made to complete the pelvic dissection via the minimally invasive robotic approach and to extract the specimen intact through a mini laparotomy. These cases were therefore classified as conversions according to the study definition, although the robotic surgical steps were completed without technical difficulty. This distinction is important. When malignancy cannot be confidently excluded preoperatively and the specimen is too large for intact vaginal extraction, oncologic principles discourage uncontained morcellation, given the risk of intraperitoneal tumor dissemination. Accordingly, conversion in these cases should be interpreted as a safety decision rather than a failure of the robotic approach. One conversion in the first phase was related to severe adhesions and uterine immobility. This supports the need for cautious selection of early cases during the first phase of a robotic program. The clinical value of maintaining safety during the learning phase should be interpreted in the broader context of robotic hysterectomy. Large systematic evidence suggests that robotic hysterectomy has perioperative outcomes largely comparable to conventional laparoscopy in routine benign cases, while it preserves the general advantages of minimally invasive surgery over open surgery, including reduced blood loss, shorter hospital stay, and fewer complications [5,23]. Therefore, the aim of robotic implementation should not be to replace laparoscopy in every patient. Rather, robotic surgery may be most useful when it helps extend minimally invasive surgery to technically demanding cases, such as patients with obesity, previous surgery, adhesions, large uterus, or complex pelvic anatomy [24–26]. Docking time deserves separate consideration. In the present study, the docking CUSUM curve reached its peak at case 28 and then progressively declined. This supports the idea that robotic setup has its own learning curve. Previous analyses of robotic draping and docking have shown that these steps improve after approximately 18–21 cases in experienced teams transitioning to a new robotic platform [27]. In our cohort, the slightly later stabilization is plausible because both the surgeon and operating room team were in their first independent robotic experience. This finding reinforces that robotic implementation is not only surgeon-dependent; it is also team-dependent. This point is also supported by recent governance literature [22]. The introduction of a robotic system affects surgeons, assistants, scrub nurses, anesthesiologists, and operating room management. Initial uncertainty, communication, docking, emergency preparedness, and system handling are common challenges during implementation. For this reason, structured training, simulation, proctoring, mentoring, and monitoring of key performance indicators are increasingly recommended when robotic programs are introduced [22]. The present study fits this framework. It provides a practical example of early institutional monitoring during the vulnerable phase of robotic adoption. The oncologic implications should also be considered with caution. Minimally invasive surgery is well established for endometrial cancer, and robotic assistance may increase the proportion of patients who can undergo minimally invasive procedures, particularly among obese, elderly, or medically complex patients [5,28,29]. In contrast, the role of minimally invasive surgery in early cervical cancer remains controversial [23,30]. For this reason, expansion toward oncologic indications should be gradual, indication-specific, and supported by careful patient counselling and outcome monitoring.

This study has several strengths. It included the first consecutive robotic hysterectomies performed by a single primary console surgeon. No cases involved console sharing. This makes the learning curve easier to interpret. The use of CUSUM analysis allowed objective visualization of performance over time. In addition, the study evaluated both operative efficiency and perioperative safety, which is essential during implementation of a new surgical program. However, several limitations should also be acknowledged. First, this was a retrospective single-center study with a limited sample size. Second, the results reflect the experience of one surgeon and one institutional team and may not be generalizable to centers with different baseline laparoscopic experience, robotic volume, training pathways, or case selection. Third, the cohort included both benign and selected malignant indications, which may introduce heterogeneity. Fourth, the learning curve was mainly assessed using time-based parameters. Operative time is useful and objective, but it does not fully capture surgical quality, tissue handling, decision-making, or long-term outcomes. Finally, the phase breakpoint was identified visually from the CUSUM curve itself and then used to test for differences between phases, an approach that carries a risk of circularity and may overstate statistical significance; formal changepoint methods (e.g., bootstrapped CUSUM or segmented regression) would offer a more robust alternative.

## 5. Conclusion

This study demonstrates that robot-assisted laparoscopic hysterectomy can be safely implemented in a tertiary university center during the initial adoption phase. Procedural proficiency was achieved after approximately 36 consecutive cases, while perioperative safety outcomes remained stable throughout the learning curve. Patients with high body mass index, large uterine volume, and low preoperative hemoglobin levels were managed safely and effectively, suggesting that these factors should not be considered barriers to the early adoption of robotic surgery within a structured training program. These findings highlight the importance of standardized training, multidisciplinary teamwork, and workflow optimization for the successful implementation of robotic gynecologic surgery.

## Declaration

### Abbreviations

CUSUM: CUmulative SUM

BMI: Body Mass Index

RAS: Robotic-assisted surgery

ASA: American Society of Anesthesiologists

### Ethics approval and consent to participate

Ethical approval was granted by the Ethics Committee Goethe University Frankfurt (Reference number 2023-1232). Informed consent was obtained from all individual participants included in the study.

### Consent for publication

Not applicable.

### Availability of data and materials

In accordance with the journal’s guidelines, we will provide our data for the reproducibility of this study if our institution approves the request.

### Author contribution

Morva Tahmasbi Rad: Supervision; Visualization; Conceptualization; Data Collection; Methodology; Formal analysis; Writing—original draft. Dario Colacurci: Investigation; Formal analysis; Writing—original draft. Elias Bascharyar: Investigation; Data collection; Formal analysis. Giuseppe Bifulco: Investigation; Data collection; Formal analysis; Sven Becker: Supervision; Conceptualization; Methodology; Writing—review & editing. Ina Shehaj: Supervision; Conceptualization; Methodology; Writing—review & editing.

## Data Availability

We will provide our data for the reproducibility of this study if our institution approves the request.

## Acknowledgements

Not applicable.

## Clinical trial number

Not applicable.

